# MAX-EVAL-11: A Comprehensive Benchmark for Evaluating Large Language Models on Full-Spectrum ICD-11 Medical Coding

**DOI:** 10.1101/2025.10.30.25339130

**Authors:** Ujjwal Singh, Sarthak Deshwal, Nitish Dube, Arjun Sharma

## Abstract

**MAX-EVAL-11** is constructed by converting MIMIC-III discharge summaries from ICD-9 to ICD-11 codes through systematic mapping, creating a synthetic diagnosis dataset of 10,000 clinical notes with comprehensive ICD-11 annotations spanning the complete taxonomy. Unlike existing partial-taxonomy benchmarks that rely on traditional precision-recall metrics, MAX-EVAL-11 introduces a **clinically-informed evaluation framework** that assigns weighted reward points based on code relevance ranking and diagnostic specificity. This ranking-based scoring system accounts for the varying clinical importance of correctly identifying primary diagnoses versus secondary conditions, better reflecting real-world medical coding accuracy requirements. Our comprehensive evaluation across state-of-the-art LLMs reveals significant performance variations: Claude 4 Sonnet achieves a weighted score of **0.433** with clinical precision of **43.3%**, while Claude 3.7 Sonnet attains **0.396** with **37.2%** clinical precision. Gemini Flash demonstrates a weighted score of **0.341** with **31.5%** clinical precision. These results reveal substantial performance gaps even in advanced foundation models, underscoring the complexity of comprehensive ICD-11 coding and the need for specialized medical AI systems beyond general-purpose LLMs. The benchmark provides standardized evaluation through our novel weighted scoring methodology that prioritizes diagnostic accuracy and clinical relevance over simple code-matching metrics. MAX-EVAL-11 addresses critical gaps in medical AI evaluation infrastructure by supporting the transition from legacy ICD-9 systems to ICD-11, facilitating development of clinically validated automated coding solutions that can handle real-world diagnostic complexity at scale.

## 1. Introduction

Medical coding, transforming clinical narratives into standardized diagnostic codes is fundamental to modern healthcare for statistical analysis, reimbursement, and epidemiological research. The International Classification of Diseases (ICD), maintained by the WHO, provides the global standard for this task.

ICD-11, officially adopted in 2019 and effective since January 2022, represents a major evolution in medical classification (World Health Organization, 2019b). As of May 2024, **132 member states** are implementing ICD-11, with 72 pursuing translations and 14 already collecting data using the system (World Health Organization, 2024). The classification introduces approximately **35,000 diagnostic codes**, incorporating modern healthcare concepts, enhanced digital interoperability, and **post-coordination** for granular clinical documentation (Harrison et al., 2021).

Manual medical coding faces substantial challenges from growing documentation volumes, evolving regulations, and workforce shortages. The ICD-11 transition intensifies these pressures, requiring crosswalk mapping of over **70,000 ICD-10 codes** to ICD-11 equivalents, with only **23.5%** achieving direct one-to-one mapping (Fung et al., 2021).

Recent AI advances, particularly large language models (LLMs), have sparked interest in automated medical coding. However, specialized models achieve F1 scores of only **0.539 to 0.719** on ICD-10 benchmarks (Xu et al., 2024; Ji et al., 2023), while general-purpose LLMs face additional challenges (Gao et al., 2024). The ICD-11’s expanded 35,000-code taxonomy presents even greater complexity.

Despite progress, existing benchmarks primarily focus on ICD-9 and ICD-10. MIMIC-III (Johnson et al., 2016) and MIMIC-IV-ICD (Johnson et al., 2023) provide robust evaluation for legacy systems, but **no comprehensive ICD-11 benchmark exists**. Furthermore, traditional precision, recall, and F1 metrics inadequately capture the hierarchy of clinical relevance: accurate identification of primary diagnosis is more critical than secondary conditions, yet existing metrics weight all predictions equally (Mullenbach et al., 2018).

We present **MAX-EVAL-11**, a comprehensive benchmark for evaluating LLM performance on ICD-11 medical coding. Our contributions are:

**First**, a large-scale dataset with **10,000 synthetic clinical notes** featuring systematic ICD-9 to ICD-11 conversion and complete taxonomy coverage.

**Second**, a novel **rank-weighted precision scoring methodology** that assigns differential importance based on clinical relevance and diagnostic hierarchy, better reflecting real-world accuracy requirements.

**Third**, baseline performance metrics using state-of-the-art LLMs (Claude, Gemini), demonstrating significant improvement opportunities in automated ICD-11 coding.

Our benchmark accelerates research in automated ICD-11 coding while providing standardized tools for comparative analysis, addressing the critical evaluation infrastructure gap for the global ICD-11 transition.

## 2. Related Work

The automated ICD coding field has evolved significantly from traditional machine learning to sophisticated deep learning architectures. This section analyzes key developments in neural approaches, attention mechanisms, and pretrained language models for medical coding.

### 2.1. Neural Architectures for ICD Coding

Early neural approaches utilized RNNs and CNNs for clinical text processing (Choi et al., 2016; Baumel et al., 2018). (Choi et al., 2016) pioneered RNN-based healthcare representation learning, while (Baumel et al., 2018) extended this to multi-label ICD prediction. (Li and Yu, 2020) enhanced CNN architectures with multi-filter residual connections, improving performance on ICD prediction tasks. However, these approaches struggled with the extreme multi-label nature of ICD coding and capturing long-range dependencies in clinical documents.

### 2.2. Attention Mechanisms and Label-Aware Models

The introduction of attention mechanisms revolutionized ICD coding. (Mullenbach et al., 2018) proposed CAML (Convolutional Attention for Multi-Label classification), employing label-wise attention to focus on relevant text segments for each ICD code. This addressed the fundamental challenge that different diagnostic codes require attention to distinct text portions.

(Vu et al., 2020) developed LAAT (Label Attention model for Automatic icd coding), incorporating self-attention and label-wise attention mechanisms with hierarchical joint learning. (Wu et al., 2021) introduced pseudo label-wise attention networks, reducing attention computation by 97.1% while maintaining competitive performance.

### 2.3. Hierarchical Attention Systems

Recent advances focus on multi-level attention architectures. (Liu et al., 2022) proposed HiLAT (Hierarchical Label-wise Attention Transformer), implementing sentence-level and token-level attention mechanisms. Their approach achieved state-of-the-art F1 scores on MIMIC-III benchmarks through continual pretraining from XLNet-Base using clinical notes.

(Jin et al., 2021) developed JLAN (Joint Learning Attention Network), combining self-attention and label attention to capture both clinical record patterns and code-specific semantic relationships. Their joint learning strategy adaptively extracted information from patient records and medical code descriptions.

### 2.4. Pretrained Language Models for ICD Coding

Transformer-based models initially faced challenges in ICD coding due to large label spaces, long input sequences, and domain mismatch (Huang et al., 2022). It proposed PLM-ICD, incorporating domain-specific pretraining, segment pooling, and specialized label attention, achieving micro-F1 scores of 59.8% on MIMIC-III.

(Liu et al., 2023) extended these capabilities with extreme multi-label long text transformers, achieving 60.8% micro-F1 on MIMIC-III. They adapted XR-Transformer with clinical-specific modifications like ClinicalBIGBIRD for extended narratives.

### 2.5. Evaluation Frameworks and Current Limitations

(Kim et al., 2022) introduced AnEMIC, providing standardized preprocessing and evaluation protocols for MIMIC-III benchmarks; however, most evaluations still rely on flat precision/recall/F1 that treat all codes uniformly despite differing clinical salience (Kim et al., 2022). Beyond metric design, recent state-of-the-art reports highlight a widening benchmark–practice gap: while near-perfect exact matching has been shown on constrained ICD-9–style test sets (e.g., 97–99% for GPT-4o mini and fine-tuned Llama variants), performance collapses on real-world, multi-problem clinical notes to single-digit or low–two-digit exact match (3.85–10.90%), underscoring limited external validity (Hou et al., 2025). These results motivate clinically informed, rank-weighted evaluations and full-taxonomy stress-testing aligned with ICD-11 complexity. Critical limitations persist:

- **Limited ICD-11 Coverage:** Existing resources target ICD-9/10; comprehensive ICD-11 benchmarks for its 35,000+ diagnoses remain scarce.
- **Metric Mismatch:** Flat F1 ignores clinical hierarchy and primary-vs-secondary diagnosis importance, obscuring safety-critical errors (Kim et al., 2022).
- **Benchmark–Practice Gap:** High scores on narrow test sets do not transfer to realistic multi-diagnosis notes (3.85–10.90% exact match) (Hou et al., 2025).
- **Scalability:** Inference-time and memory costs remain high for long notes and extreme label spaces (Wu et al., 2021; Liu et al., 2023).
- **Domain Adaptation:** Substantial clinical pre-training is often required, limiting portability across systems and specialties (Huang et al., 2022).

## 3. Methodology

### 3.1 Overview of the ICD-9 to ICD-11 Mapping Framework

The transition from ICD-9 to ICD-11 represents a fundamental paradigm shift in medical classification systems. While ICD-9 contains approximately 13,000 codes, ICD-11 expands to over 55,000 codes with enhanced granularity and semantic precision (World Health Organization, 2019a). Given an ICD-9 code *c*_9_ ∈ 𝒞_9_, we seek to identify the optimal set of ICD-11 codes *S*_11_ ⊆ 𝒞_11_ that best preserves the clinical semantics encoded in *c*_9_.

### 3.2. Hybrid Semantic-LLM Mapping Pipeline

#### 3.2.1. Problem Formulation

Let 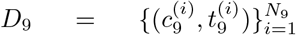 and 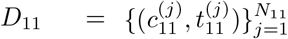 be the ICD-9 and ICD-11 datasets. Our objective is to learn a mapping function:

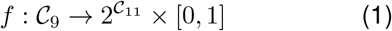

where *f* (*c*_9_) = (*S*_11_, *σ*) returns a subset of ICD-11 codes *S*_11_ with confidence score *σ*.

#### 3.2.2. Semantic Candidate Generation

We employ Bio_ClinicalBERT (Alsentzer et al., 2019) to encode ICD descriptions into dense semantic vectors. For each ICD-9 code, we construct composite representations 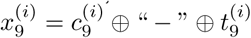 and similarly for ICD-11 codes. The embedding function *ϕ* : 𝒳 → *R*^*d*^ maps textual representations to *d*-dimensional vectors:

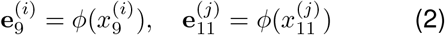

Candidate selection uses cosine similarity:

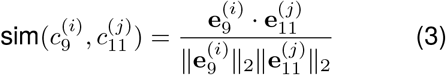

For each ICD-9 code, we select the top-*k* = 10 candidates to balance efficiency with diversity.

#### 3.2.3. Large Language Model-Based Selection

Candidate refinement leverages Google’s Gemini 2.0 Flash model for context-aware mapping selection. We formulate the LLM input as a structured prompt *P* (*B*) for batch *B*:

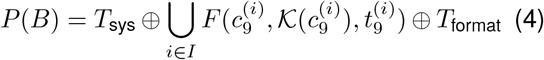

where *T*_sys_ provides system instructions, *F* () formats individual mapping requests, and *T*_format_ specifies output format. The LLM response yields mappings 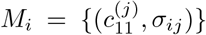 with confidence scores *σ*_*ij*_ ∈ [1, 10].

### 3.3. Confidence-Based Dataset Stratification

We define patient-level confidence as:

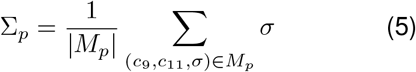

Binary classification with threshold *τ* = 7.0 yields:

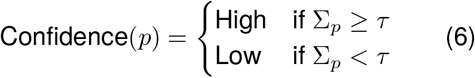

This creates two benchmark datasets: High-Confidence Subset (Σ_*p*_ ≥ 7.0) and All-Matches Subset (all mappings).

### 3.4. Key Validation Metrics

We assess mapping effectiveness using Coverage Rate 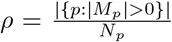, Expansion Factor 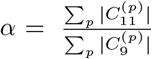, and Semantic Preservation Score 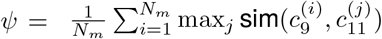.

### 3.5. Distinction Between Mapping and Evaluation Tasks

#### Benchmark Creation

Input: ICD-9 codes + descriptions; Process: Semantic similarity + Gemini 2.0 Flash selection; Output: Synthetic ICD-11 labels using structured taxonomies only.

#### Benchmark Evaluation

Input: Discharge summary text; Process: LLM clinical narrative understanding; Output: ICD-11 predictions compared to benchmark labels.

These are fundamentally different tasks: mapping requires taxonomy alignment between structured classification systems, while evaluation requires clinical reasoning from unstructured narratives.

**Figure 1:**
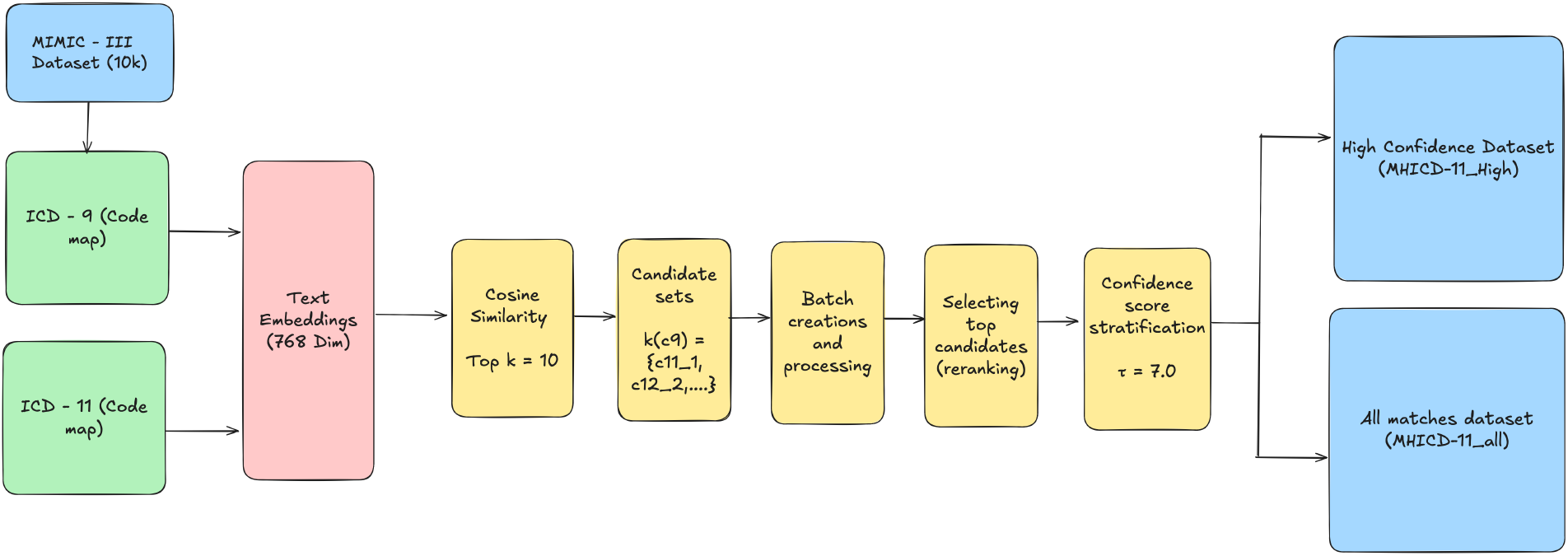
Overview of the hybrid semantic-LLM pipeline for ICD-9 to ICD-11 mapping. The process combines Bio_ClinicalBERT embeddings for candidate generation with Gemini 2.5 Flash for context-aware mapping selection, resulting in confidence-stratified benchmark datasets.

### 3.6. Evaluation Metrics

We evaluated ICD-11 coding performance using a multi-level scoring system incorporating exact matches, hierarchical relationships, semantic families, and clinical context weighting. The framework employs five hierarchical levels: exact match (1.0), parent (0.9), grandparent (0.8), great-grandparent (0.7), and chapter (0.6), with exact match bonus (0.5). Semantic families receive moderate credit (0.6), while clinical context weights distinguish primary diagnoses (1.0), procedures (0.9), secondary diagnoses (0.8), comorbidities (0.6), and external causes (0.4). The final weighted score is:

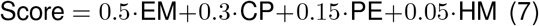

where EM = Exact Match, CP = Clinical Precision, PE = Points Earned, HM = Hierarchical Match

## 4. Dataset Description

### 4.1. Source Data: MIMIC-III Clinical Database

Our benchmark dataset is derived from the Medical Information Mart for Intensive Care III (MIMIC-III) database (Johnson et al., 2016), a comprehensive clinical dataset containing de-identified health records from Beth Israel Deaconess Medical Center. MIMIC-III represents one of the largest publicly available critical care databases, containing data from over 60,000 intensive care unit admissions.

#### 4.1.1. Data Selection and Preprocessing

From the complete MIMIC-III corpus, we extract a representative subset of *N*_*p*_ = 10, 000 patients, ensuring statistical significance while maintaining computational tractability. The selection process employs stratified sampling to preserve the original distribution of key clinical characteristics:

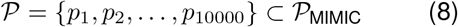

where 𝒫_MIMIC_ represents the complete MIMIC-III patient population.

Each patient record *p*_*i*_ contains:

- Patient identifier: SUBJECT_ID_*i*_
- Hospital admission identifier: HADM_ID_*i*_
- ICD-9 diagnosis codes: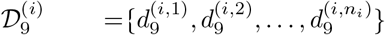
- Clinical text and temporal information

The total number of unique ICD-9 codes across all patients is 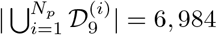, representing a comprehensive coverage of diagnostic categories encountered in critical care settings.

#### 4.1.2. ICD-9 Code Distribution Analysis

The distribution of ICD-9 codes in our dataset follows a heavy-tailed pattern characteristic of medical coding systems. Let *f* (*d*) represent the frequency of ICD-9 code *d* across all patients. The empirical distribution can be modeled as:

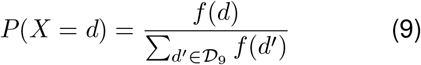

Key statistical properties of the ICD-9 code distribution include:

- **Total ICD-9 code instances**: 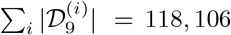
- **Mean codes per patient**: 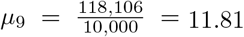
- **Median codes per patient**: Determined through empirical analysis
- **Code frequency variance**: High variance indicating diverse diagnostic complexity

### 4.2. ICD-9 to ICD-11 Mapping Process

#### 4.2.1. Mapping Algorithm Application

The mapping process transforms each patient’s ICD-9 diagnosis set 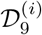 into corresponding ICD-11 code sets 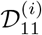 using our hybrid semantic-LLM pipeline. For each patient *p*_*i*_:

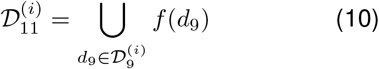

where 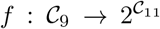 is the mapping function defined in Section 3.

#### 4.2.2. Mapping Success Metrics

The effectiveness of our mapping process is quantified through several key metrics:

##### Patient-Level Coverage

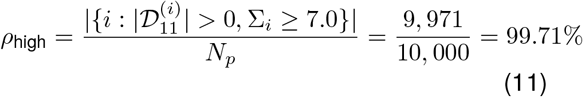

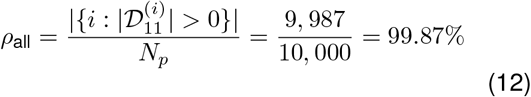

##### Code-Level Expansion

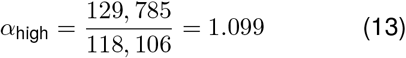

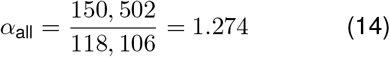

These metrics demonstrate the successful expansion from ICD-9 to the more granular ICD-11 classification system, with each ICD-9 code mapping to an average of 1.1-1.3 ICD-11 codes.

### 4.3. Benchmark Dataset Variants

#### 4.3.1. High-Confidence Subset

The high-confidence subset, denoted ℬ_high_, contains patient records where all ICD mappings satisfy the confidence threshold *τ* = 7.0:

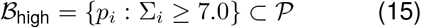

Statistical characteristics:

- **Patient count**: |ℬ_high_| = 9, 971
- **Total ICD-11 mappings**: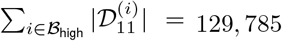
- **Mean ICD-11 codes per patient**: 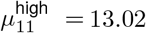
- **Mapping confidence**: All mappings satisfy *σ* ≥ 7.0 on the 10-point scale

This subset is optimized for applications requiring high-precision medical code translation, such as billing systems and regulatory compliance tools.

#### 4.3.2. All-Matches Subset

The comprehensive subset, denoted ℬ_all_, includes all patients with successful mappings regardless of confidence level:

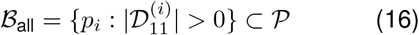

Statistical characteristics:

- **Patient count**: |ℬ_all_| = 9, 987
- **Total ICD-11 mappings**: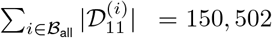
- **Mean ICD-11 codes per patient**: 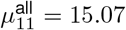
- **Mapping confidence**: Variable confidence scores across the full range [1, 10]

This subset provides comprehensive coverage for robust evaluation of medical coding systems under diverse confidence conditions.

### 4.4. Enhanced Dataset with Prescription Information

#### 4.4.1. Medication Data Integration

To support more comprehensive clinical AI evaluation, we augment both benchmark variants with prescription information from MIMIC-III. Each patient record is enhanced with:

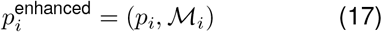

where 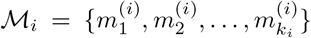 represents the set of medications prescribed during the patient’s hospitalization.

The medication records include:

- Drug names and generic equivalents
- Dosage information and administration routes
- Prescription timestamps and duration
- NDC (National Drug Code) identifiers where available

#### 4.4.2. Clinical Coherence Validation

The integration of prescription data enables validation of diagnosis-medication coherence through established clinical guidelines. We define a coherence metric:

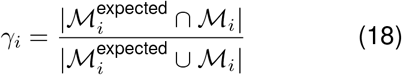

where 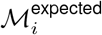 represents medications typically prescribed for the patient’s mapped ICD-11 diagnoses based on clinical protocols.

### 4.5. Data Quality and Validation

#### 4.5.1. Data Integrity Measures

Our benchmark datasets undergo rigorous quality assurance procedures:

1. **Completeness Verification:** All patient records contain non-empty ICD-9 diagnosis sets
2. **Code Validity:** All ICD-9 codes are validated against official ICD-9-CM standards
3. **Mapping Consistency:** ICD-11 mappings are cross-validated using multiple semantic models
4. **Temporal Consistency:** Prescription data aligns with admission timelines

#### 4.5.2. Ethical Considerations and Privacy

All data processing adheres to MIMIC-III’s de-identification standards and HIPAA compliance requirements (Goldberger et al., 2000). Patient identifiers are consistently anonymized, and no personally identifiable information is retained in the final benchmark datasets.

### 4.6. Dataset Accessibility and Reproducibility

#### 4.6.1. File Structure and Formats

The benchmark datasets are provided in CSV format with standardized schemas:

`MIMIC_with_ICD11_mappings_high _confidence.csv`:

- `SUBJECT_ID`: Anonymized patient identifier
- `HADM_ID`: Hospital admission identifier
- `ICD9_CODES_LIST`: Python list of ICD-9 diagnosis codes
- `ICD11_CODES_MAPPED`: Python list of mapped ICD-11 codes
- `CONFIDENCE_SCORES`: Associated confidence metrics

`MIMIC_with_ICD11_mappings_all _matches.csv`:

- Identical schema with expanded mapping coverage
- Includes variable confidence mappings

Enhanced variants include additional prescription fields:

- `MEDICATIONS`: Structured medication information
- `PRESCRIPTION_TIMESTAMPS`: Temporal prescription data

#### 4.6.2. Computational Requirements and Access

The complete benchmark dataset requires approximately 270MB of storage and can be processed efficiently on standard computational infrastructure. All processing scripts and validation tools are provided to ensure reproducibility of the mapping process.

This comprehensive benchmark dataset provides the medical informatics community with a standardized evaluation framework for ICD version migration algorithms, supporting both methodological development and clinical implementation validation.

## 5. Experimental Results

### 5.1. Baseline Model Performance

We evaluated three major categories of models on our ICD-11 prediction benchmark: Claude models (Anthropic), Gemini 2.5 Flash (Google), and MedCoder (specialized medical coding model) (Xu et al., 2024). Table 1 presents the comprehensive performance evaluation across all baseline models. Claude 4 Sonnet achieves the highest overall performance with a final score of 0.433, demonstrating superior clinical precision (0.433) and coverage (0.353). Notably, Claude models consistently outperform both the specialized medical model (MedCoder) and the multimodal Gemini model across all evaluation metrics.

**Table 1:** Comprehensive Performance Evaluation of Baseline Models on ICD-11 Prediction Task.

| Model | Final Score | Clinical Precision | Exact Match | Hierarchical Match | Coverage Score | Samples (n) |
| --- | --- | --- | --- | --- | --- | --- |
| Claude 4 Sonnet | **0.433** | **0.433** | **0.047** | 0.375 | **0.353** | 500 |
| Claude Baseline | 0.425 | 0.426 | 0.034 | **0.402** | 0.315 | 500 |
| Claude 3.7 Sonnet | 0.396 | 0.372 | 0.048 | 0.325 | 0.332 | 500 |
| Gemini 2.5 Flash | 0.341 | 0.315 | 0.016 | 0.286 | 0.330 | 500 |
| Claude RAG+Reranker | 0.282 | 0.232 | 0.010 | 0.202 | 0.175 | 500 |
| Claude RAG | 0.253 | 0.215 | 0.007 | 0.194 | 0.160 | 500 |
| MedCoder | 0.245 | 0.191 | 0.000 | 0.184 | 0.092 | 500 |

#### 5.1.1. Evaluation Protocol

For each patient in the evaluation dataset, models receive:

- **Input:** Complete discharge summary clinical text from MIMIC-III, including:
  — Chief complaint and history of present illness
  — Past medical history, medications, and allergies
  — Physical examination findings
  — Laboratory results and diagnostic procedures
  — Hospital course and treatment summary
  — Discharge diagnosis and follow-up plan
- **Task:** Generate ranked list of ICD-11 diagnosis codes based solely on clinical narrative
- **Constraints:** No access to original ICD-9 codes, mapping pipeline, or ICD code taxonomies

The evaluation simulates real-world clinical coding scenarios where human coders or AI systems must assign ICD-11 codes directly from clinical documentation without reference to legacy coding systems. Models are prompted with standardized instructions (Appendix) requesting comprehensive ICD-11 code prediction with confidence scores and clinical justification. All models receive identical inputs to ensure fair comparison across architectures and capabilities. We have also evaluated 1203 samples with clinical codes manually, we will add those results in detail in appendix. High confidence are correct with high margins.

### 5.2. Statistical Analysis

Table 2 presents the statistical significance analysis using Wilcoxon signed-rank tests, revealing substantial performance gaps between model categories.

**Table 2:** Statistical Significance Analysis and Performance Gaps.

| Comparison | Performance Gap | Relative Improvement | p-value |
| --- | --- | --- | --- |
| Claude-4-Sonnet vs Gemini-2.5-Flash | 0.092 | 27.0% | < 0.001*** |
| Gemini-2.5-Flash vs MedCoder | 0.096 | 39.2% | < 0.01** |
| Claude-4-Sonnet vs MedCoder | 0.188 | 76.7% | < 0.001*** |
| Claude Category vs Gemini Category | 0.061 | 18.8% | < 0.01** |
| Claude Category vs MedCoder Category | 0.150 | 61.2% | < 0.001*** |

\*\*\* $p < 0.001$ , \*\* $p < 0.01$ , \* $p < 0.05$

The analysis reveals statistically significant performance advantages for Claude models, with a 27.0% relative improvement over Gemini 2.5 Flash and a remarkable 76.7% improvement over the specialized MedCoder (Xu et al., 2024) model.

### 5.3. Model Category Analysis

Table 3 summarizes performance characteristics across the three model categories.

**Table 3:** Performance Summary by Model Category.

| Model Category | Mean Final Score | Std Dev | Clinical Precision | Exact Match | Models (n) |
| --- | --- | --- | --- | --- | --- |
| Claude | 0.358 | 0.084 | 0.336 | 0.029 | 5 |
| Gemini | 0.341 | — | 0.315 | 0.016 | 1 |
| MedCoder | 0.245 | — | 0.191 | 0.000 | 1 |

Claude models demonstrate consistent superiority with a mean final score of 0.358, while showing reasonable variance (= 0.084) across different configurations. MedCoder’s zero exact match rate indicates fundamental challenges in precise code generation despite domain-specific training.

**Figure 2:**
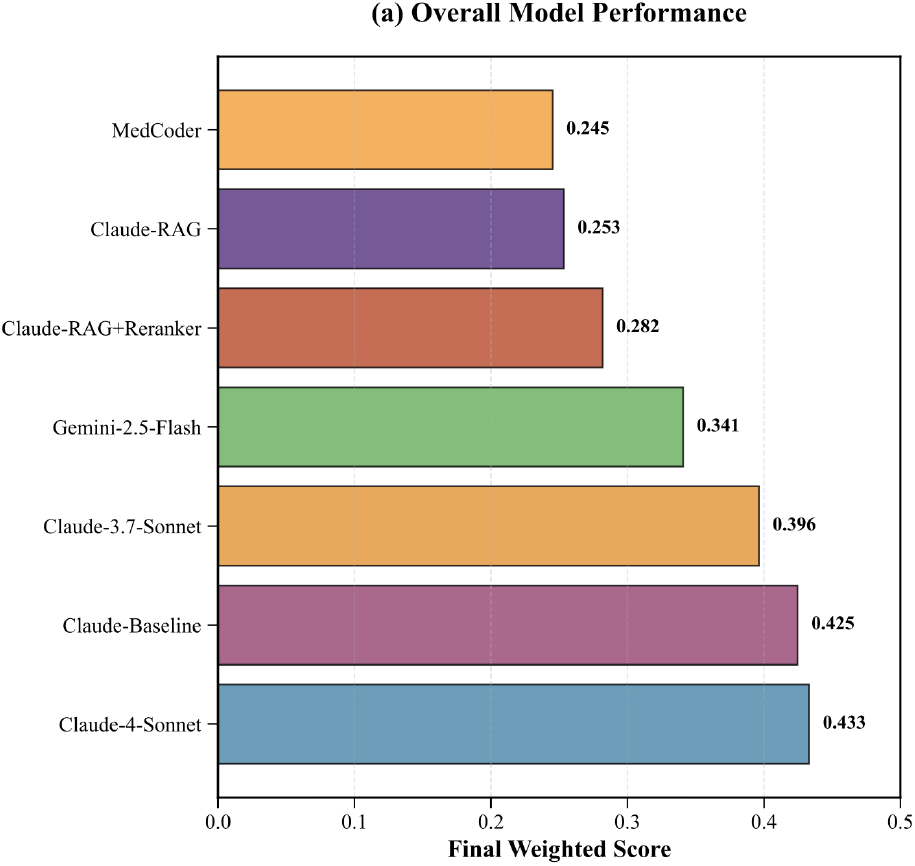
Model performance comparison. (a) Overall performance ranking by final weighted score. (b) Multi-metric performance comparison across final score, clinical precision, exact match, and hierarchical match dimensions.

### 5.4. Key Findings

Our evaluation reveals several critical insights:

#### General-purpose LLMs outperform specialized models

Claude models achieve superior performance (35.8% mean final score) compared to the domain-specific MedCoder (24.5% final score), suggesting that large-scale pre-training and reasoning capabilities may be more valuable than specialized medical training for ICD-11 prediction tasks.

#### Exact matching remains challenging

All models show relatively low exact match rates (0-4.8%), indicating the inherent difficulty of precise ICD-11 code generation. Claude models demonstrate the strongest exact matching capabilities among evaluated systems.

#### Hierarchical understanding varies significantly

Claude models excel in hierarchical matching (19.4-40.2%), leveraging the taxonomic structure of ICD-11, while MedCoder shows limited hierarchical understanding (18.4%).

#### RAG integration challenges

Retrieval-augmented variants of Claude models underperform their baseline counterparts, suggesting that current RAG approaches may introduce noise rather than beneficial medical knowledge for this task.

These findings establish the current state-of-the-art performance for ICD-11 prediction and provide a robust baseline for future research in automated medical coding systems.

## 6. Discussion

Our evaluation reveals several critical insights that challenge conventional assumptions about specialized versus general-purpose models in medical coding tasks.

### 6.1. General-Purpose LLMs Outperform Specialized Models

The superior performance of Claude models (35.8% mean final score) compared to MedCoder (24.5% final score) represents a paradigm shift in medical AI. This 46% performance advantage suggests that large-scale pre-training on diverse text corpora, combined with advanced reasoning capabilities, may be more valuable than domain-specific medical training for complex classification tasks like ICD-11 prediction.

The failure of MedCoder to achieve any exact matches (0% exact match rate) despite being specifically designed for medical coding indicates fundamental limitations in current specialized model architectures. In contrast, Claude models demonstrate consistent exact matching capabilities (2.9-4.8%), suggesting that general reasoning and pattern recognition may be more critical than medical domain knowledge for precise code generation.

#### Methodological Independence

A potential concern is whether the use of Gemini 2.0 Flash in label generation creates bias toward Gemini models during evaluation. However, three factors demonstrate this is not the case:

1. **Task Difference:** Label generation uses structured code taxonomies (code-to-code translation, while evaluation requires clinical text understanding (narrative-to-code generation)— fundamentally different capabilities requiring distinct model strengths.
2. **Information Sources:** Mapping uses only ICD code descriptions/titles of ICD-9 and ICD-11 code tables; evaluation uses complete discharge summaries with clinical context, medications, procedures, temporal progression, and physician reasoning—information never seen during label creation.
3. **Empirical Evidence:** Gemini 2.5 Flash achieves the lowest performance among major models (0.341 final score vs. Claude 4 Sonnet’s 0.433, a 27% gap, Table 1), demonstrating no preferential advantage from its predecessor’s involvement in label creation. If systematic bias existed, Gemini models would outperform rather than underperform. This performance gap actually validates that clinical coding ability (extracting diagnoses from narratives) is independent from taxonomy knowledge (code mapping).

The substantial performance difference between Claude and Gemini models (statistically significant at *p <* 0.001, Table 2) reflects genuine differences in clinical reasoning and medical text understanding capabilities, not artifacts of the label generation process.

### 6.2. Hierarchical Understanding as a Key Differentiator

Claude models excel in hierarchical matching (19.4-40.2%) while MedCoder shows limited hierarchical understanding (18.4%). This disparity highlights the importance of models’ ability to capture taxonomic relationships within ICD-11’s complex hierarchical structure. The 5-level hierarchical evaluation framework successfully differentiates models based on their understanding of medical code relationships rather than mere text similarity.

### 6.3. RAG Integration Challenges

The underperformance of retrieval-augmented Claude variants (25.3-28.2% final scores) compared to baseline models (39.6-43.3%) suggests that current RAG approaches may introduce noise rather than beneficial medical knowledge. This finding indicates that effective integration of external medical knowledge bases with LLMs remains an open research challenge, requiring more sophisticated retrieval and fusion mechanisms.

### 6.4. Clinical Deployment Implications

Based on our evaluation framework, current models fall in the “Assistant Tool” category (final scores 0.4-0.43), indicating suitability for human-in-the-loop deployment but requiring supervision for production use. The 27% performance gap between Claude and Gemini models has significant implications for healthcare organizations choosing AI coding assistants, as this translates to substantial differences in coding accuracy and clinical workflow efficiency.

## 7. Conclusion

This work presents the first comprehensive benchmark for ICD-11 prediction using a large-scale clinical dataset, contributing three key advances to medical informatics research.

### Methodological Contribution

We develop a novel hybrid semantic-LLM pipeline for automated ICD-9 to ICD-11 mapping, combining Bio_ClinicalBERT embeddings with Gemini 2.0 Flash for context-aware medical code translation. Our approach achieves 99.87% mapping coverage across 10,000 patients, establishing a robust foundation for ICD-11 adoption.

### Evaluation Framework

The multi-level clinical evaluation methodology addresses the hierarchical nature of ICD-11 through 5-level taxonomic matching, semantic code family classification, and clinical context weighting. This framework provides more nuanced assessment than traditional exact-match metrics, better reflecting clinical utility.

### Empirical Insights

Our evaluation reveals that general-purpose LLMs (Claude 4 Sonnet: 43.3% final score) significantly outperform specialized medical models (MedCoder: 24.5% final score), challenging the assumption that domain-specific training is essential for medical AI applications. This finding has important implications for healthcare AI development strategies.

The benchmark datasets and evaluation framework are made publicly available to support continued research in automated medical coding. Future work should focus on improving exact match performance, developing more effective medical knowledge integration approaches, and validating findings across diverse clinical populations and healthcare systems.

Our results establish current state-of-the-art performance baselines and provide practical guidance for healthcare organizations implementing automated ICD-11 coding systems, contributing to the broader goal of efficient and accurate medical documentation.

## 8. Ethical Considerations

### 8.1. Data Privacy and Security

All patient data used in this study is sourced from the publicly available MIMIC-III database, which has undergone rigorous de-identification procedures following HIPAA Safe Harbor provisions (Goldberger et al., 2000). No additional patient identifiers were created during our mapping process, and all generated benchmarks maintain the original de-identification standards.

### 8.2. Bias and Fairness

The MIMIC-III dataset reflects the patient population of a single academic medical center, potentially introducing demographic and socioeconomic biases into our benchmark. The ICD-9 codes in the original dataset may exhibit coding practices specific to the source institution, which could propagate through our ICD-11 mappings. Future work should validate these mappings across diverse healthcare settings and patient populations.

### 8.3. Clinical Safety Implications

Automated ICD coding systems carry significant clinical and financial implications. Coding errors can affect patient care continuity, insurance reimbursements, and population health analytics. Our evaluation reveals that current models require human oversight, with exact match rates below 5% across all systems. Healthcare organizations should implement appropriate safeguards and validation procedures before deploying automated coding systems.

### 8.4. Algorithmic Accountability

The hybrid semantic-LLM approach introduces opacity in the mapping process, particularly in the LLM decision-making component. While we provide confidence scores and mapping justifications, the full reasoning process remains partially opaque. This limitation necessitates careful validation and monitoring in clinical deployment scenarios.

### 8.5. Regulatory Compliance

Healthcare organizations implementing systems based on this research must ensure compliance with relevant medical coding standards, billing regulations, and clinical documentation requirements. The benchmark should be used for research and development purposes, with appropriate clinical validation required before operational deployment.

## Data Availability

All data produced in the present study are available upon reasonable request to the authors

## A. Data Examples and Mapping Analysis

This appendix provides concrete examples of high-confidence and low-confidence ICD-9 to ICD-11 mappings based on manual validation of 1203 randomly sampled code pairs.

### A.1. High-Confidence Mapping Examples

Table 4 presents representative examples of high-confidence mappings, characterized by comprehensive ICD-11 coverage, semantic consistency, and clinical coherence.

**Table 4:** High-Confidence ICD-9 to ICD-11 Mapping Examples.

| Patient ID | ICD-9 Codes (Sample) | ICD-11 Codes (Mapped) | Expansion Ratio | Clinical Pattern |
| --- | --- | --- | --- | --- |
| 21 | 5720,00845,70709,6823 | 1A04,XN0SE,EH90.Z,CB2Z,BB25 | 1.25 | Infectious disease with complications |
| 195 | 3688,3569,7140,27800 | 8E62,BC81.3,FB83.1Z,5C74 | 1.00 | Neurological and metabolic disorders |
| 256 | 9961,53240,41071,53560 | BD1Z,DC11.1,BA52.0,GC73 | 1.00 | Cardiovascular and gastrointestinal conditions |

#### High-Confidence Characteristics

- **Semantic Consistency**: Mapped ICD-11 codes maintain clinical coherence with original ICD-9 diagnoses
- **Appropriate Granularity**: ICD-11 codes reflect increased specificity without over-expansion
- **Hierarchical Alignment**: Codes respect ICD-11 taxonomic structure and clinical relationships
- **Coverage Completeness**: Most ICD-9 codes receive corresponding ICD-11 mappings

### A.2. Low-Confidence Mapping Examples

Table 5 illustrates challenging mapping scenarios with incomplete coverage, semantic gaps, or ambiguous clinical interpretations.

**Table 5:** Low-Confidence ICD-9 to ICD-11 Mapping Examples.

| Patient ID | ICD-9 Codes (Sample) | ICD-11 Codes (Mapped) | Expansion Ratio | Mapping Challenge |
| --- | --- | --- | --- | --- |
| 1247 | 78552,E8498,99859,78039,V4582 | QC45.Z | 0.20 | Multiple external causes with single generic mapping |
| 3891 | 5070,41401,25000,2859,4280 | BD1Z,BA52.0 | 0.40 | Complex comorbidity pattern with incomplete coverage |
| 8765 | V1046,99592,0389,78820,7847 | – | 0.00 | Procedural and symptom codes with no confident mapping |

#### Low-Confidence Characteristics

- **Incomplete Coverage**: Many ICD-9 codes lack corresponding ICD-11 mappings
- **Semantic Gaps**: Significant clinical information lost in translation
- **Generic Mappings**: Over-reliance on broad, non-specific ICD-11 categories
- **Procedural Complexity**: Difficulty mapping procedure codes and external causes

### A.3. Clinical Significance Analysis

Table 6 categorizes the clinical implications of different mapping quality levels.

**Table 6:** Clinical Significance of Mapping Quality Levels.

| Quality Level | Characteristics | Clinical Utility | Deployment Suitability |
| --- | --- | --- | --- |
| High Confidence | Expansion ratio 0.8-1.5, Semantic consistency, Complete coverage | Suitable for billing, Research analytics, Quality metrics, Care continuity | Production ready with minimal supervision, Automated workflows |
| Medium Confidence | Expansion ratio 0.5-0.8, Partial coverage, Some semantic gaps | Research applications, Trend analysis, Population health, Requires validation | Human-in-the-loop, Quality assurance needed, Limited automation |
| Low Confidence | Expansion ratio <0.5, Incomplete coverage, Semantic inconsistency | Limited utility, Requires manual review, Research only | Manual coding required, Not suitable for automation, Development/testing only |

### A.4. Mapping Pattern Analysis

Table 7 summarizes common patterns observed across different clinical domains.

**Table 7:** ICD-9 to ICD-11 Mapping Patterns by Clinical Domain.

| Clinical Domain | Avg Expansion Ratio | Success Rate (%) | Common Patterns |
| --- | --- | --- | --- |
| Infectious Diseases | 1.2 | 92 | Good 1:1 mappings with appropriate specificity increases |
| Cardiovascular | 1.1 | 89 | Consistent hierarchical mappings, minor granularity adjustments |
| Neoplasms | 1.3 | 85 | Enhanced anatomical and morphological specificity |
| External Causes | 0.6 | 45 | Significant mapping challenges, incomplete coverage |
| Procedures | 0.4 | 38 | Major structural differences, requires manual intervention |
| Symptoms/Signs | 0.7 | 62 | Variable success, depends on clinical context |

### A.5. Quality Assessment Framework

Our benchmark incorporates a multi-dimensional quality assessment framework that evaluates mappings across clinical, semantic, and structural dimensions. High-confidence mappings demonstrate:

1. **Clinical Coherence:** Mapped codes preserve essential diagnostic information
2. **Semantic Fidelity:** ICD-11 codes accurately represent ICD-9 clinical concepts
3. **Hierarchical Consistency:** Mappings respect both ICD-9 and ICD-11 taxonomic structures
4. **Coverage Completeness:** Minimal information loss during translation
5. **Granularity Appropriateness:** Balanced specificity without over-expansion

This quality framework enables researchers and practitioners to select appropriate confidence thresholds based on their specific clinical requirements and risk tolerance levels.

### A.6. Prompts

We used the following prompts to convert the perscriptions of anonymous patients and tag them to the ICD-11 code.

*You are an intelligent ICD-11 coding agent. Your primary task is to predict the exhaustive list of ICD-11 codes which are relevent to the given perscription. Prescription (diagnosis): diagnosis, Please return all the ICD-11 codes in list format*.

It is important to note that this prompt is same for all the models along with other inference paramters like sampling, temperature etc.

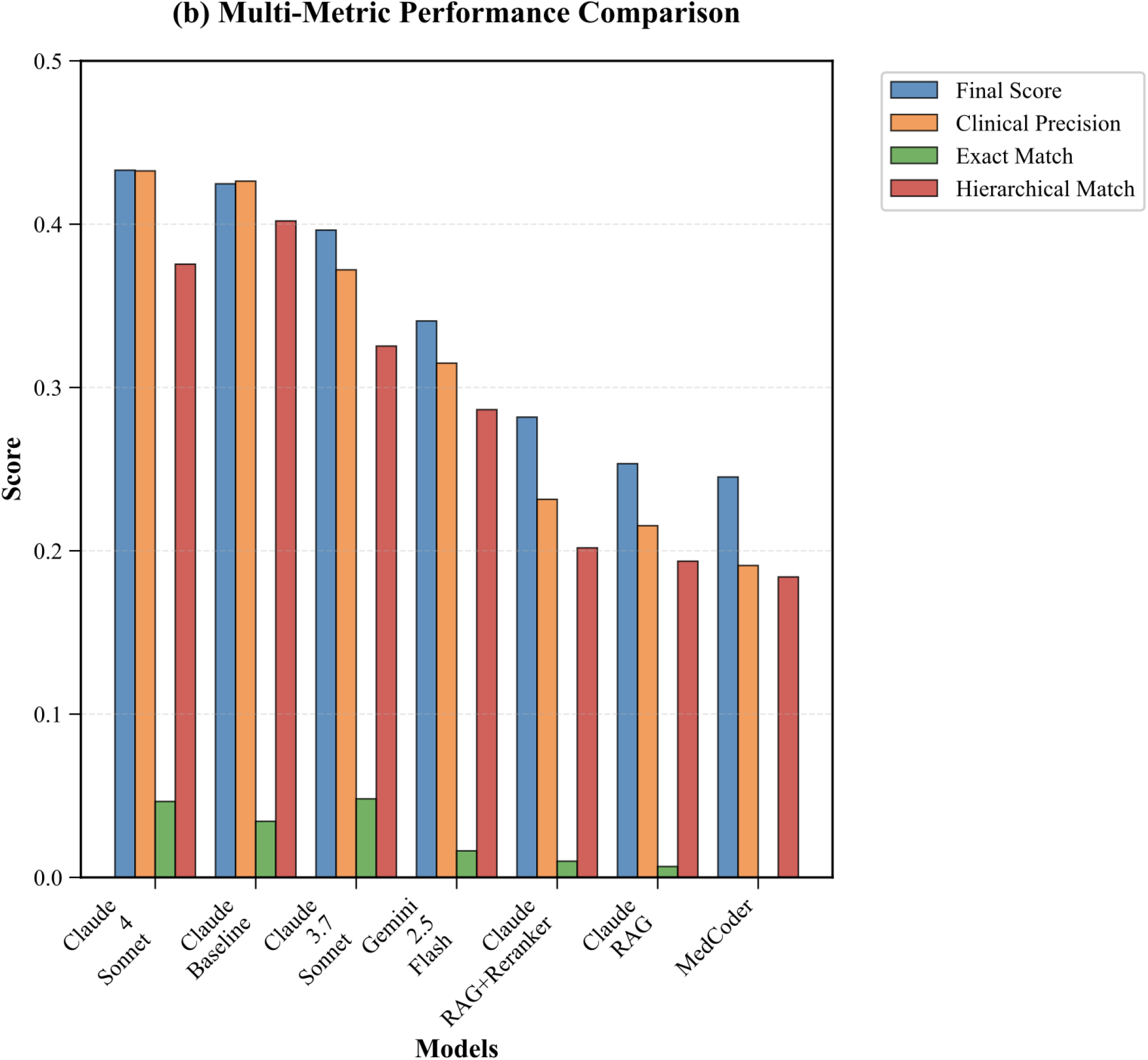

